# Knowledge is not all you need to generate trust in AI use in healthcare

**DOI:** 10.1101/2024.01.10.24301097

**Authors:** Anson Kwok Choi Li, Ijaz A. Rauf, Karim Keshavjee

## Abstract

**Background:** Canada has invested significantly in artificial intelligence (AI) research and development over the last several years. Canadians’ knowledge of and attitudes towards AI in healthcare are understudied.

**Objectives:** To explore the relationships between age, gender, education level, and income on Canadians’ knowledge of AI, their comfort with its use in healthcare, and their comfort with using personal health data in AI research.

**Methods:** Ordinal logistics regression and multivariate polynomial regression were applied to data from the 2021 Canadian Digital Health Survey using RStudio and SigmaZone’s Design of Experiments Pro.

**Results:** Female and older Canadians self-report less knowledge about AI than males and other genders and younger Canadians. Female Canadians and healthcare professionals are less comfortable with use of AI in healthcare compared to males and people with other levels of education. Discomfort appears to stem from concerns about data security and the current maturity level of the technology.

**Conclusion:** Knowledge of AI and the use of AI in healthcare are inversely correlated with age and directly correlated with education and income levels. Overall, female respondents self-reported less knowledge and comfort with AI in healthcare and research than other genders. Privacy concerns should continue to be addressed as a major consideration when implementing AI tools. Canadians, especially older females, not only need more education about AI in healthcare, but also need more reassurance about the safe and responsible use of their data and how bias and other issues with AI are being addressed.

**Author Summary:** Artificial intelligence (AI) and its application has garnered significant public interest and excitement within healthcare in recent years. However, its successful integration and use in healthcare will depend on patient and user adoption. As a result, AI tools may be limited in healthcare when user concerns are not carefully addressed and if patients are not educated about how these technologies work. While there have been studies on the attitudes of clinicians and healthcare professionals toward AI, little is known about the general public’s perception of AI within the healthcare setting. Our study addresses this gap in the literature by analyzing data from the 2021 Canadian Digital Health Survey to understand the relationships between Canadians’ attitudes towards AI and various socioeconomic and demographic factors. Our results found that older Canadians, Canadians with less formal education and women need to be better informed about the safe and responsible use of AI and be reassured about good data security practices before it can be broadly accepted by them. In addition, the element of trust may be a factor that is contributing to the higher levels of discomfort with AI observed in middle-aged Canadians. The findings from this study will help stakeholders better implement and broaden the accessibility of AI technologies.

## Introduction

Artificial intelligence (AI) has garnered significant public interest in recent years as it has become increasingly more integrated into everyday life. The global AI healthcare market was valued at US $11.2 billion in 2022 and is estimated to achieve a market size of US $427.5 billion by 2032, an over 40-fold increase over just a decade [1]. Canada has a large AI community and research hub and the country is ranked 1st among G7 nations in the five-year average year-over-year growth rate in AI talent concentration [2]. Canada was the first country to launch a national AI strategy and has earmarked $443.8 million to help with the commercialization and adoption of AI technology over 10 years starting in 2021 [3].

The application of AI within healthcare has received significant global interest and enthusiasm in recent years. Its application is already widespread and spans different areas, such as drug discovery [4], oncology [5], anesthesiology [6], and dentistry [7]. The benefits of implementing AI in healthcare are substantial, both changing the way medicine is practiced and reducing healthcare costs [8,9]. AI tools have the potential to help patients stay healthy through earlier diagnosis, tailored treatments, and better monitoring, which are likely to lead to fewer hospitalizations and decreased use of the healthcare system. There have been multiple studies on the attitudes of clinicians and healthcare professionals toward AI, however, there is a dearth of studies related to the general public’s perception of AI [10]. Canadians have a generally positive outlook on the impact of AI on their lives, but very little is known about their attitudes toward AI tools within the healthcare setting.

A recent study using the 2021 Canadian Digital Health Survey looked at the perceptions of AI in healthcare among younger (under 55 years of age) and older (over 55 years of age) Canadians and found that older Canadians are more open to the applications of AI within healthcare compared to younger Canadians [12]. While age may be an important factor in understanding Canadians’ attitudes towards AI, other socioeconomic and demographic factors may also play a role. As a result, it is important to understand the attitudes and perceptions of a range of Canadians towards AI technologies and how those attitudes differ based on various social and demographic factors before developing ways to optimize patient adoption of AI healthcare tools.

Successful adoption of healthcare technologies depends on patient acceptance of those technologies [13]. When patients don’t understand how a technology works, are unsure of its benefits or have doubts about the technology, they are less likely to accept them in their care. Without careful attention to educating those less knowledgeable and addressing important concerns of average citizens and those from vulnerable and marginalized populations, we may witness an AI divide, just as we have experienced a digital divide over the last decade concerning mobile and digital technologies.

Trust is the lubricant of any healthcare system. Patients trust their doctors to keep their information private and make decisions in their best interests. Trust enables researchers to access de-identified data for research. Trust enables double-blind research to be conducted. Trust is required to use data for training and operationalizing AI in healthcare.

Trust can be strained or even broken when patients are not consulted or they hear about data breaches or flaws in the execution of newer technologies. There are many assaults on trust in the healthcare system today including, the potential for data breaches and identity theft, misinformation, algorithmic bias, black box AI and data misappropriation.

When trust is not established or is lost, patients hesitate to participate. Australia’s personally controlled electronic health record system (PCEHR) is a case in point. In 2013, Australia embarked on the PCEHR project, touting the benefits of a patient-controlled electronic health record system. However, the government failed to get physicians on board. Physicians felt that having a patient-controlled EHR could compromise the completeness of data in the EHR. Patients got mixed messages from their physicians and the government and lost trust in the system. They also did not participate. It was only when the government re-established trust with both parties was the project able to move forward [14].

As the Canadian government continues to invest heavily in AI research and works towards positioning the country as a major AI hub, it is crucial to understand how AI technologies can be implemented and successfully adopted by end users in the healthcare setting, especially amongst historically marginalized populations and those who have traditionally been left behind.

In this paper, we analyze data from the 2021 Canadian Digital Health Survey commissioned by Canada Health Infoway, focusing on questions about the knowledge and comfort a range of Canadians (based on age, gender, education and income) have with using AI in healthcare. We identified gaps in knowledge and comfort that Canadians have with the use of AI in healthcare and make recommendations on how to address them.

## Methods

### Recruitment and Data Collection

Data was obtained from the 2021 Canadian Digital Health Survey, a cross-sectional survey of 12,052 Canadians over 16 years of age. It was commissioned by Canada Health Infoway and conducted by Leger via computer-assisted web interviewing technology (CAWI). The data was collected between July 14 to August 6, 2021.

The Canadian Digital Health Survey collected data on attitudes, utilization, perceptions, and expectations of digital health services in Canada. Survey participants were selected from the Leger Opinion panel, a group of 500,000 representative panelists from all regions of Canada. The panelists were randomly selected, and participants from hard-to-reach target groups were added to the panel through targeted recruitment campaigns. The survey was available to the respondents in either English or French [15].

### Measures

The following questions were used to assess the attitudes and comfort level of respondents towards the use of AI in the healthcare setting:

- Q.40 -On a scale of 1-4 with 1 being not at all knowledgeable and 4 being very knowledgeable, How knowledgeable are you about what artificial intelligence is?
- Q.41 -On a scale of 1-4 with 1 being very uncomfortable and 4 being very comfortable: AI has major potential when used in healthcare because it can rapidly process vast amounts of information. How comfortable are you with AI being used as a tool in healthcare?
- Q.42 -How comfortable are you with scientists using personal health data for AI research as long as informed consent has been provided by the patient?
- Q.43 -How comfortable are you with scientists using personal health data for AI research without informed consent as long as it is deidentified (i.e., personal information such as name, date of birth has been removed)?

### Independent Variable

The four independent variables used in this study included age, gender, highest level of education attained, and household income (Table 1). Age was calculated as the difference between a respondent’s birth year and the survey date.

**Table 1.** Descriptive statistics of respondents based on socioeconomic and demographic characteristics.

| Factor | Age Category (Code) | Sample Size<br>(Male - 1) | Sample Size<br>(Female - 2) | Sample Size<br>(Other - 3) |
| --- | --- | --- | --- | --- |
| Age | 16-24 (1) | 274 | 549 | 28 |
|  | 25-34 (2) | 676 | 802 | 18 |
|  | 35-54 (3) | 870 | 815 | 19 |
|  | 55-64 (4) | 983 | 851 | 12 |
|  | 65+ (5) | 1841 | 1444 | 16 |
| Income | < \$24,999 | 321 | 455 | 26 |
| | \$25,000-\$49,999 | 737 | 871 | 21 |
| | \$50,000-\$79,999 | 1035 | 1044 | 18 |
| | \$80,000-\$99,000 | 804 | 685 | 7 |
| | \$100,000-\$149,999 | 1054 | 860 | 13 |
| | \$150,000-\$249,999 | 556 | 457 | 5 |
| | \$250,000+ | 137 | 89 | 3 |
| Education | Highschool | 827 | 975 | 37 |
|  | Apprenticeship/Trades | 323 | 185 | 2 |
|  | College/CEGEP | 1027 | 1195 | 17 |
|  | University degree | 1716 | 1578 | 23 |
|  | Masters | 597 | 432 | 11 |
|  | PhD | 105 | 66 | 0 |
|  | Medical/paramedical | 49 | 30 | 3 |

Data points were excluded from the analyses if a respondent answered “Prefer not to answer” for the household income question and/or “Other,” “None of the above,” or “Prefer not to answer” for the highest level of education obtained question.

### Outcome (Dependent) Variable

The dependent variables included the respondent’s self-reported knowledge of artificial intelligence and comfort levels with artificial intelligence and its implications on personal health data used in healthcare.

Data points were not included in the analyses if a respondent answered “Don’t know” for Q.41, Q42 and Q43 where “Don’t know” was an option.

### Statistical Analysis

#### Design

This study employed ordinal logistic regression models to investigate the associations between dependent and independent variables, including age, income, education level, and gender. The dependent variables were transformed and ranked on an ordinal scale from lowest (1) to highest (4), reflecting the different response options. Multivariate polynomial regression was performed to gain deeper insights into the underlying relationships, considering both linearity and quadratic effects and exploring potential interactions between different variables influencing responses.

#### Data Transformation

Options for each dependent variable were systematically transformed into ordinal categories, and the independent variables (age, income, education level, and gender) were encoded as factors. This transformation allowed for the application of ordinal logistic regression models, capturing the ordered nature of the dependent variable responses.

#### Software and Tools

Ordinal logistic regressions were conducted using RStudio, a comprehensive, integrated development environment for R. Ordinal logistic regression models were fitted to explore the relationships between the dependent and independent variables. The models aimed to assess the impact of age, income, education level, and gender on the ordered response categories.

Multivariate polynomial regression was conducted using the Design of Experiments (DOE) Pro (obtained from SigmaZone) software addon within Microsoft Excel. The multivariate polynomial regression allowed a more nuanced exploration of the complex interactions and non-linear associations between variables.

Collectively, these analytical approaches provide a comprehensive understanding of the relationships between the variables under investigation, offering insights into both linear and non-linear patterns and potential interactions influencing the ordinal responses.

## Results

### Descriptive Statistics

Descriptive data of the respondents’ sociodemographic characteristics and responses to Q40-Q43 are shown in Table 1.

We report on 5 key findings:

1. Women of all ages self-report less knowledge about AI than their male and Other counterparts (Figure 1). This finding was consistent across all income and educational levels.
2. Older Canadians, regardless of gender, income or education, self-report less knowledge about AI than younger and middle-aged Canadians (Figure 1). More educated Canadians reported being more knowledgeable about AI than less educated Canadians.
3. Middle-aged and older Canadian women are the least *comfortable* with the use of AI in healthcare (Figure 2). The non-linear relationships demonstrated in Figure 2 in contrast to the linear relationships in Figure 1 tell us that comfort with the use of AI does not correlate highly with knowledge of AI.
4. Discomfort with AI correlates highly with the use of consented (identifiable) data in research (Figure 3), but not with the use of anonymized (de-identified) data (Figure 4).
5. Paradoxically, healthcare professionals expressed less comfort with the use of AI than other educated Canadians (Table 2).

**Fig 1.**
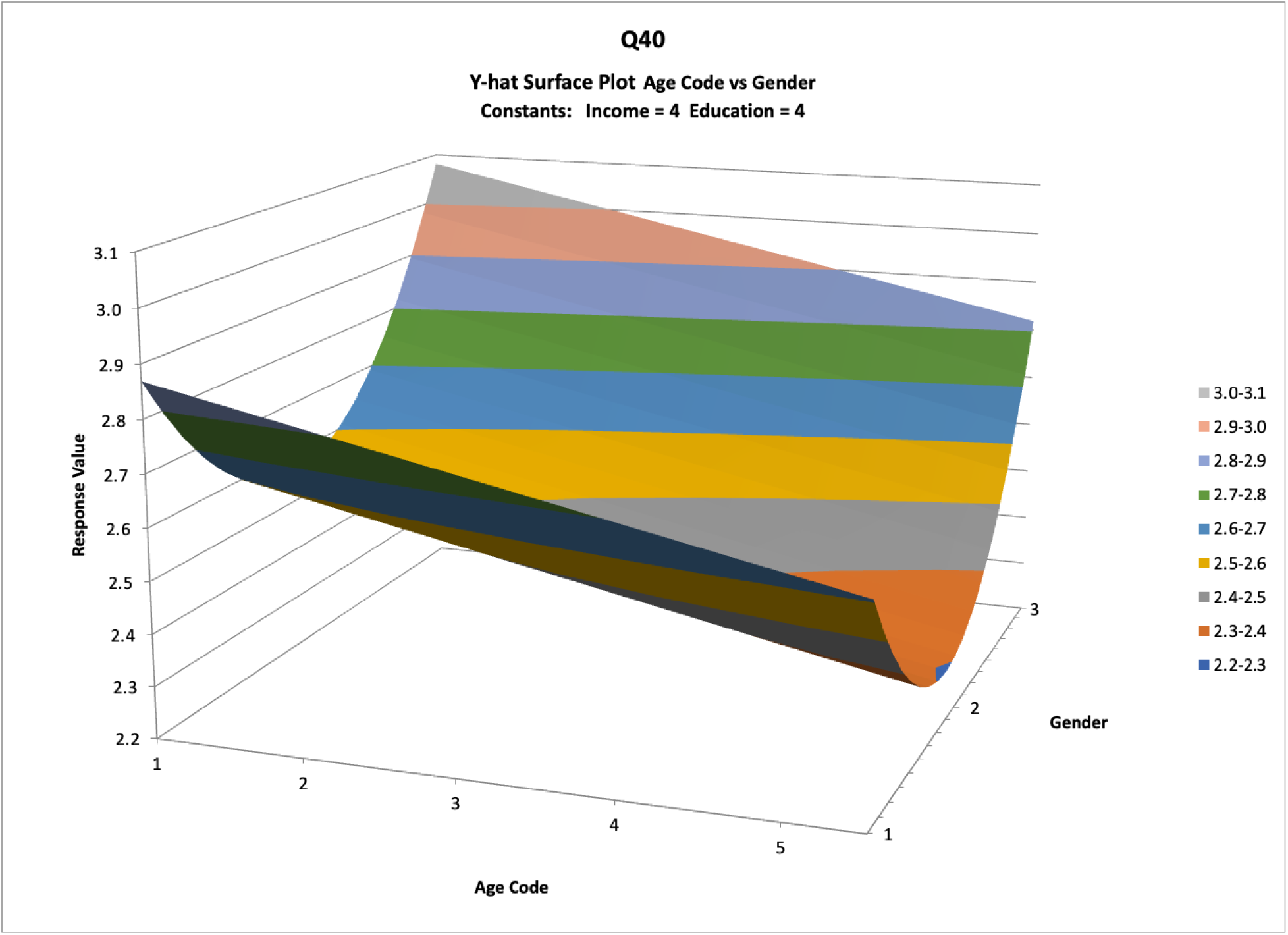
Pareto chart of the ŷ values of the respondents’ knowledge of AI against age and gender. (Age Code) 1 = 16-24, 2 = 25-34, 3 = 35-54, 4 = 55-64, 5 = 65+. (Gender) 1= Male, 2 = Female, 3 = Other.

**Fig 2.**
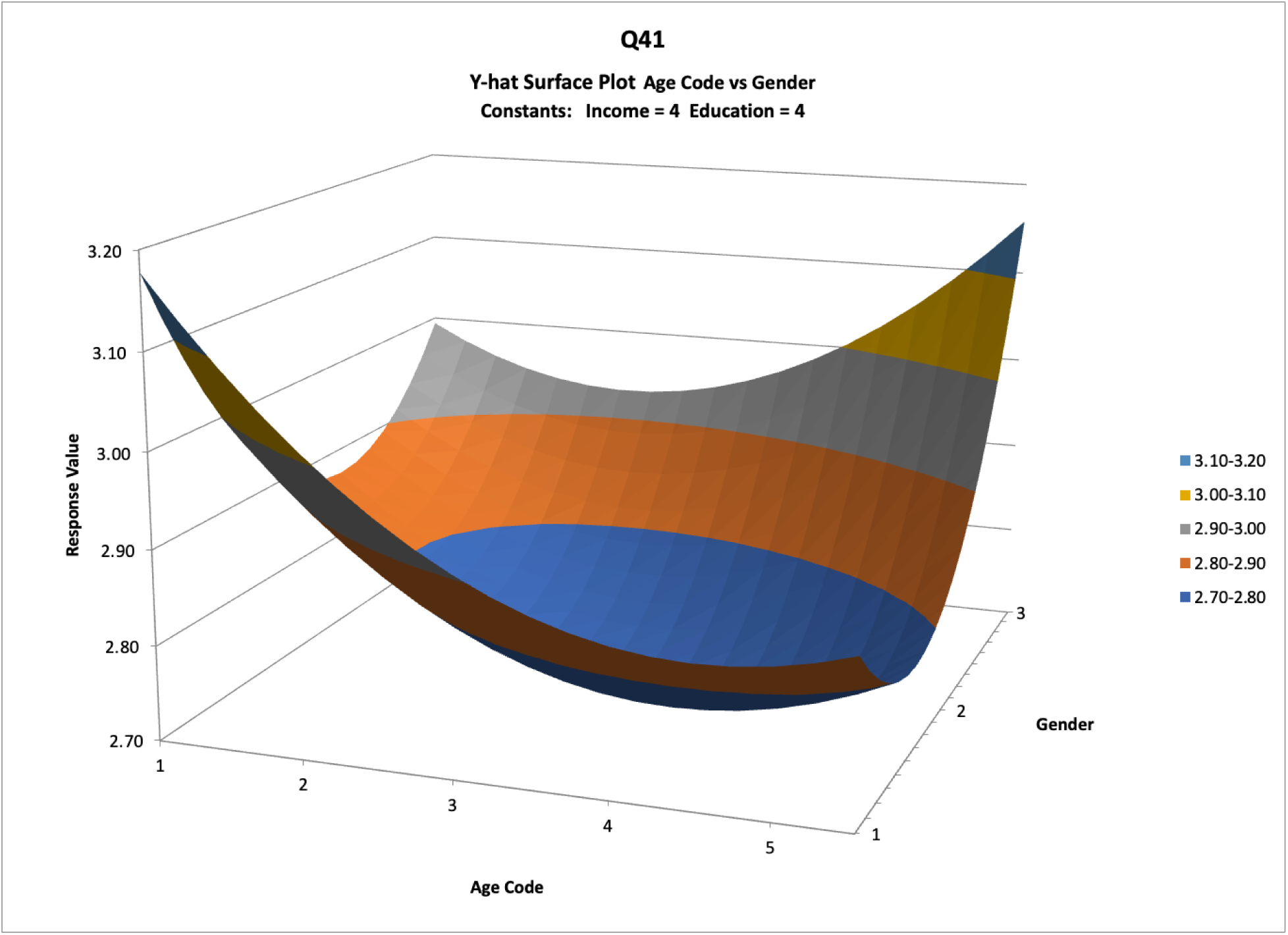
Pareto chart of the ŷ values of the respondents’ comfort with AI being used in healthcare against age and gender. (Age Code) 1 = 16-24, 2 = 25-34, 3 = 35-54, 4 = 55-64, 5 = 65+. (Gender) 1= Male, 2 = Female, 3 = Other.

**Fig 3.**
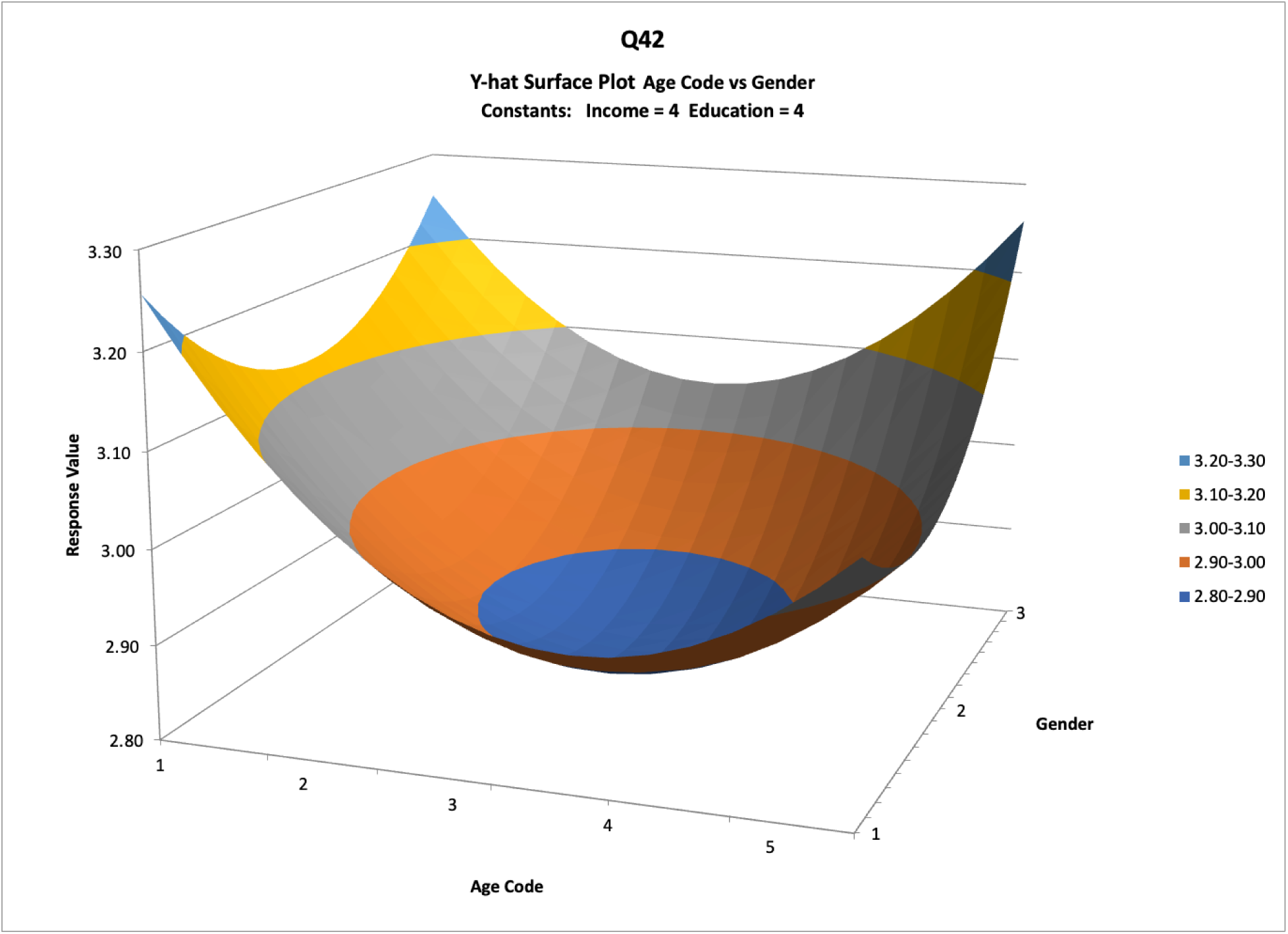
Pareto chart of the ŷ values of the respondents’ comfort with consented data being used for AI research against age and gender. (Age Code) 1 = 16-24, 2 = 25-34, 3 = 35-54, 4 = 55-64, 5 = 65+. (Gender) 1= Male, 2 = Female, 3 = Other.

**Fig 4.**
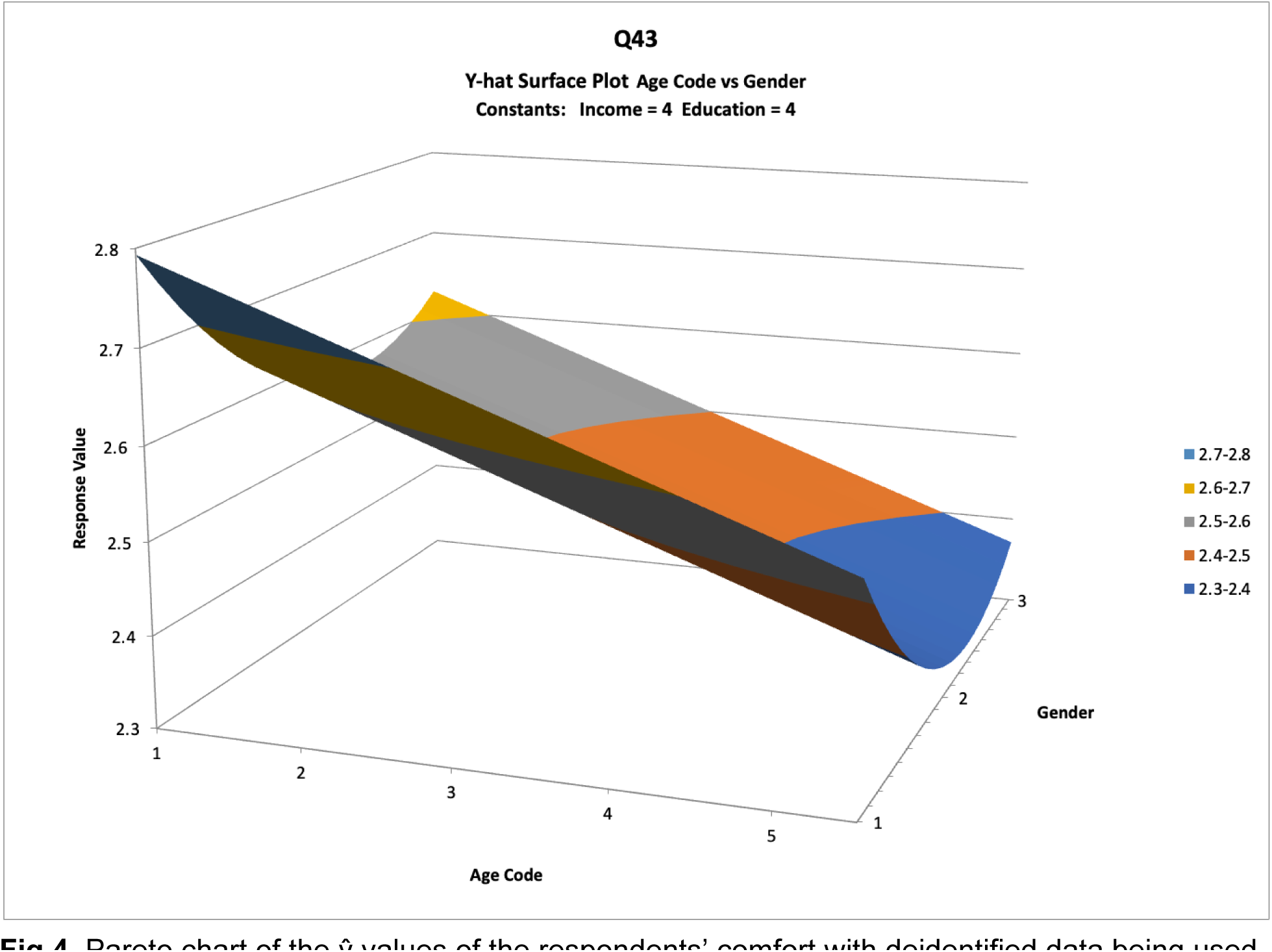
Pareto chart of the ŷ values of the respondents’ comfort with deidentified data being used for AI research against age and gender. (Age Code) 1 = 16-24, 2 = 25-34, 3 = 35-54, 4 = 55-64, 5 = 65+. (Gender) 1= Male, 2 = Female, 3 = Other.

**Table 2.**
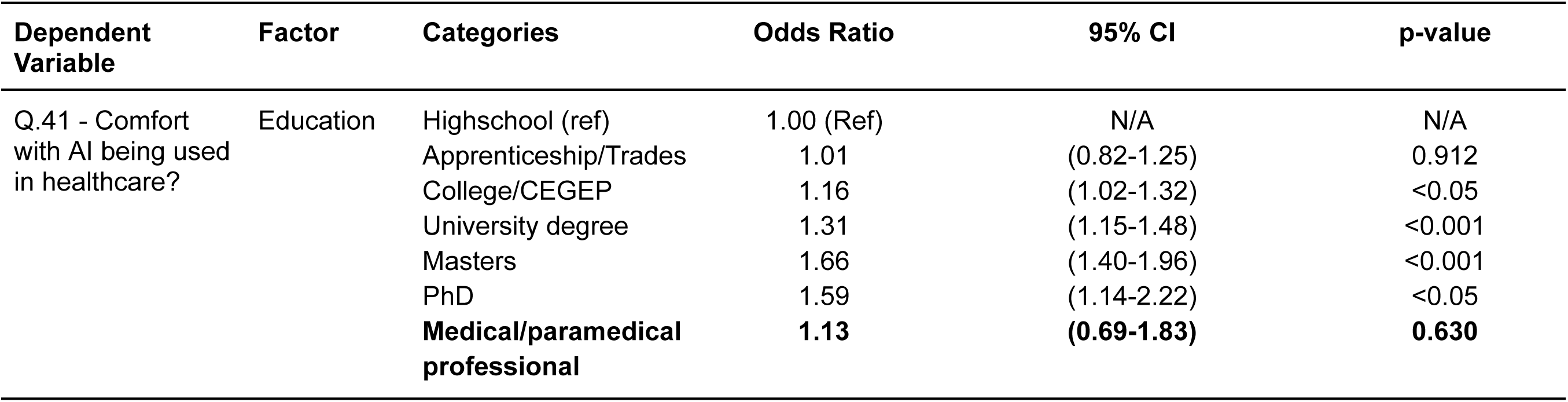
Likelihood of being comfortable with AI compared to reference group.

| **Dependent Variable** | **Factor** | **Categories** | **Odds Ratio** | **95% CI** | **p-value** |
| --- | --- | --- | --- | --- | --- |
| Q.41 - Comfort with AI being used in healthcare? | Education | Highschool (ref) | 1.00 (Ref) | N/A | N/A |
|  |  | Apprenticeship/Trades | 1.01 | (0.82-1.25) | 0.912 |
|  |  | College/CEGEP | 1.16 | (1.02-1.32) | <0.05 |
|  |  | University degree | 1.31 | (1.15-1.48) | <0.001 |
|  |  | Masters | 1.66 | (1.40-1.96) | <0.001 |
|  |  | PhD | 1.59 | (1.14-2.22) | <0.05 |
|  |  | **Medical/paramedical professional** | **1.13** | **(0.69-1.83)** | **0.630** |

## Discussion

The findings from this study demonstrate that attitudes towards AI in healthcare and research are impacted by the complex relationships between various socioeconomic and demographic factors, including gender, age, income, and education. The study also demonstrates that knowledge of AI is not sufficient to generate comfort with the use of AI in healthcare or the conduct of research using identifiable data, even if it is consented. While there has been significant attention and excitement with the application of AI technology in healthcare, our findings suggest that not all Canadians are equally receptive to its use.

The knowledge and comfort levels of AI use in healthcare and research were significantly lower between the respondents who identified as female and those who identified as male/Other across all dependent variables. Due to the self-reporting nature of the CDHS, it is not possible to discern whether female respondents truly have less knowledge, are underestimating their knowledge, or if the male and ‘Other’ respondents are overestimating their knowledge or a combination of the three. Previous research has found that women are underrepresented in the AI sector in Canada [16]. This gender gap within the AI research community may also extend to the general public, generating the findings we observed in this study.

It appears that knowledge of AI is not sufficient to explain the levels of discomfort that some Canadians, especially middle-aged women, express with the use of AI in healthcare. The non-linear relationship between comfort with AI and age (Fig 2) suggests that other factors are likely to be at play. While the CDHS only collected data on knowledge and comfort with the use of AI, trust may be another factor that explains the deviation seen in the middle-aged group. The stark difference in response to the question about research using identifiable vs. de-identified data makes it clear that the identifiability of data is concerning to middle-aged individuals, especially women. Deidentification of data removes the element of trust compared to consented data and as a result, may be more correlated with knowledge than comfort. This is reflected in the linear relationship found between age and both knowledge of AI and comfort with deidentified data being used for research, where no deviation in the middle-aged group was observed. The discomfort with identifiable data research may be driven by the many data breaches experienced by healthcare organizations in recent years and widely reported in the media. Data breaches and the risk of identity theft may be driving concerns about and discomfort with AI use among middle-aged Canadians.

While older Canadians contribute the largest proportion of healthcare utilization within Canada [17], the middle-aged population’s level of trust may be due to a less positive perception of the healthcare system compared to the other age groups as a result of their interactions with different parts of the healthcare system, for example, primary care, which is under significant stress. Previous studies in Canada have found variability in patient attitudes toward consent preferences of health data being used for research purposes [18].

Interestingly, healthcare professionals also expressed discomfort with the use of AI in healthcare. Given that most healthcare professionals likely understand the value of consented (identifiable) research, it is unlikely that their discomfort is driven only by concerns about data safety. Other factors may also be at play, including concerns about the maturity of the technology, algorithmic bias, and AI explainability. Limitations of our study include self-report bias since this study uses the data from the Canadian Digital Health Survey, a self-reported survey. The validity and reliability of the data cannot be determined due to the nature of the survey design. Given the general nature of the questions in the survey, the interpretation of the question depends on the respondent; different respondents may interpret the question very differently. However, given the large and varied sample, the results provide relevant insight into Canadians’ attitudes toward the use of AI in healthcare and research.

## Conclusion and Recommendations

Despite the significant research that has been performed on AI tools and applications thus far in healthcare, there is a lack of data on Canadians’ attitudes toward AI in general and how various socioeconomic and demographic factors affect them. Understanding the factors that influence attitudes from the perspective of the target user of these tools is important for stakeholders involved in AI research, development, and commercialization.

This study found several correlations between age, gender, income level, and education level and attitudes towards the use of AI in healthcare and research.

Older Canadians, Canadians with less formal education and women need to be better informed about AI in general before it will be broadly accepted by these groups. However, knowledge about AI will not be enough. Creating trust requires good governance and good communication. The healthcare system not only needs good governance, it needs to be seen to be well governed. Understanding how trust impacts the middle-aged group regarding AI use in healthcare and consented data used in AI research should be explored in future research. Multiple trust factors at play need to be teased out, including concerns about data breaches, identity theft, misinformation, misappropriation of data, algorithmic bias, and unexplainable AI, amongst others.

More needs to be done to reduce the gender gap in AI and specifically to increase female representation in the AI space. Addressing this social gap between female and male/Other respondents may help to improve overall comfort with consented/identifiable data use in AI research and the use of AI in healthcare in general.

Healthcare providers have unique insider knowledge and their discomfort with the use of AI in healthcare should be explored in greater depth. What do they know that others don’t know? Will their concerns evaporate with increased maturity of the technology or are there significant underlying issues that need to be addressed? This also is a topic for future research.

The findings from this study will help AI developers, policymakers, clinicians, and other stakeholders to get started with addressing key gaps in knowledge and comfort in the use of AI in healthcare and help identify where further research is required. This will ultimately help them to develop ways to optimize the approach for the successful adoption and integration of AI tools within healthcare and research.

Understanding Canadians’ priorities and preferences with personal health data will help improve the overall acceptance of its use in AI research. Despite the enthusiasm that many Canadians have for the application of AI in healthcare, consent, and privacy of health data is still a consideration and concern that they share. The Canadian healthcare system needs to increase its protection of patient data and demonstrate that data security and safety are priorities.

## Data Availability

The public-use dataset file for the 2021 Canadian Digital Health Survey was accessed using the Borealis data online tool. The file and further information can be obtained here: https://borealisdata.ca/dataverse/canadahealthinfoway

## Author Contributions

- Anson Kwok Choi Li: Conceptualization, Data Curation, Formal Analysis, Investigation, Methodology, Validation, Writing - Original Draft Preparation, and Writing - Review & Editing

- Ijaz A. Rauf: Conceptualization, Formal Analysis, Investigation, Methodology, Validation, Visualization, Writing - Original Draft Preparation, and Writing - Review & Editing

- Karim Keshavjee: Conceptualization, Investigation, Methodology, Supervision, Validation, Writing - Original Draft Preparation, and Writing - Review & Editing

